# The association between cardiometabolic index and rheumatoid arthritis in American adults: the potential mediating role of hypertension, diabetes, and inflammatory markers

**DOI:** 10.1101/2025.10.08.25337615

**Authors:** Keju Wang, Xuefei Li, Hao Wan, Peng Tian, Chang Lu, Xiaolin Ding

**Author notes:** KW and XL are joint first authors on this work.

## Abstract

**Introduction:** The cardiometabolic index (CMI) is a novel integrated measure of dyslipidemia (as reflected by the triglyceride to high-density lipoprotein cholesterol ratio) and central adiposity (as represented by the waist-to-height ratio). While CMI has been linked to various metabolic disorders, its association with rheumatoid arthritis (RA) remains unclear. This study aimed to examine the relationship between CMI and RA among U.S. adults and to investigate potential mediating pathways.

**Methods:** We analyzed data from 9,765 adults participating in the National Health and Nutrition Examination Survey (NHANES) between 2007 and 2018. Weighted multivariable logistic regression models were used to evaluate the association between CMI and RA. Nonlinearity was assessed using restricted cubic splines (RCS). Subgroup analyses were performed to test the robustness of the findings, and mediation analysis was conducted to quantify the contributions of hypertension, diabetes, and inflammatory markers.

**Results:** In fully adjusted models, each unit increase in CMI was positively associated with RA risk (OR = 1.20, 95% CI: 1.08–1.33).. Participants in the highest CMI tertile had a 74% increased risk of RA compared to those in the lowest tertile (OR = 1.74, 95% CI: 1.33–2.28). RCS analysis identified a nonlinear relationship with a threshold effect at a CMI value of 1.63. Below this inflection point, the association was more pronounced (OR = 1.53, 95% CI: 1.27–1.85). Exploratory mediation analyses showed that hypertension (explaining 22.2% of the total association) and diabetes (15.9%) statistically accounted for a significant proportion of the association, with neutrophil count also contributing modestly (5.4%).

**Conclusions:** Higher CMI is positively associated with RA prevalence among U.S. adults, with evidence of a threshold effect. This relationship may be largely statistically explained by metabolic comorbidities,, particularly hypertension and diabetes.

## Introduction

Rheumatoid arthritis (RA) is a chronic inflammatory disorder primarily characterized by joint inflammation and pain [1]. Its global prevalence is estimated to range between 0.24% and 1.25%, with significant variation across geographic regions and ethnic groups [2]. Beyond imposing a substantial burden on individual health and quality of life, RA also contributes considerably to societal and economic costs [3]. With the growing global disease burden, the number of individuals affected by RA is projected to reach approximately 31.7 million by 2050 [4]. The disease not only causes progressive joint damage and functional impairment but is also associated with an increased risk of cardiovascular disease, malignancy, and chronic kidney disease [5, 6]. Hence, early intervention, lifestyle adaptation, and preventive strategies are essential for mitigating disease risk and improving long-term outcomes.

The pathogenesis of RA involves a complex interplay of immune cell activation, release of inflammatory mediators, and metabolic dysregulation [7]. Conventional perspectives emphasize genetic susceptibility and intrinsic immune dysregulation—particularly aberrant activation of T and B lymphocytes and amplified pro-inflammatory cytokine cascades—as central to RA development [8, 9]. However, recent advances in the field of immunometabolism have reshaped our understanding of RA pathogenesis [10]. Emerging evidence indicates that immune cell functionality is intimately linked to metabolic reprogramming, whereby shifts in metabolic pathways directly influence immune cell activation, proliferation, and effector responses. For instance, effector T cells rely on a metabolic switch from oxidative phosphorylation to glycolysis to support their activation and clonal expansion. Dysregulation of this process is now recognized as a key driver of RA pathology [11–13]. Notably, such cellular metabolic alterations are closely tied to systemic metabolic health. Epidemiological studies have established that metabolic disturbances—including obesity, insulin resistance, and dyslipidemia—are independent risk factors for RA, contributing to both its prevalence and progression [14–16]. Central obesity, defined by excessive visceral adipose tissue accumulation, is of particular relevance. Visceral fat functions not only as an energy reservoir but also as an active endocrine organ, secreting pro-inflammatory adipokines (e.g., leptin, resistin) and cytokines (e.g., TNF-*α*, IL-6), which are known mediators in RA pathogenesis [17]. Similarly, atherogenic dyslipidemia—marked by high triglycerides and low HDL-C—is not only a hallmark of cardiovascular disease but also promotes immune dysfunction and exacerbates chronic inflammation [18].

The cardiometabolic index (CMI), which integrates measures of atherogenic dyslipidemia (TG/HDL-C ratio) and central obesity (waist-to-height ratio), has recently gained attention as a composite marker in chronic disease research [19, 20]. Elevated CMI has been positively associated with the incidence of type 2 diabetes, coronary heart disease, and stroke [21–23], and serves as a reliable predictor of cardiometabolic risk. It also correlates with increased inflammatory activity and disease severity in obese and metabolically compromised individuals [24]. Despite this, most studies on CMI have focused on metabolic and cardiovascular conditions, with its potential role in rheumatoid arthritis remaining unexplored.

This study utilizes data from the U.S. National Health and Nutrition Examination Survey (NHANES) spanning 2007 to 2018 to examine the association between the CMI and RA among U.S. adults.We hypothesized that higher CMI levels would be independently associated with an increased prevalence of RA, and that this association would be partly explained by metabolic comorbidities (hypertension and diabetes) and systemic inflammation.

## Materials and methods

### Study design and population

The National Health and Nutrition Examination Survey (NHANES), conducted by the National Center for Health Statistics (NCHS), is designed to evaluate the health and nutritional status of the non-institutionalized U.S. population. The survey employs a complex, multistage probability sampling design. A multidisciplinary research team, comprising physicians, health technicians, and interviewers, administered standardized interviews and physical examinations to all participants. Participants received compensation for their involvement. This cross-sectional analysis initially included 59,842 participants from seven NHANES cycles spanning 2007 to 2018. We excluded individuals aged *<*20 years (*n* = 25, 072), those with missing data on arthritis status or the cardiometabolic index (CMI) (*n* = 20, 457), participants with other forms of arthritis (*n* = 2, 568), and those with incomplete data on potential mediators or covariates (*n* = 1, 980). After these exclusions, 9,765 participants were retained for the final analysis (Figure1).

**Fig 1.**
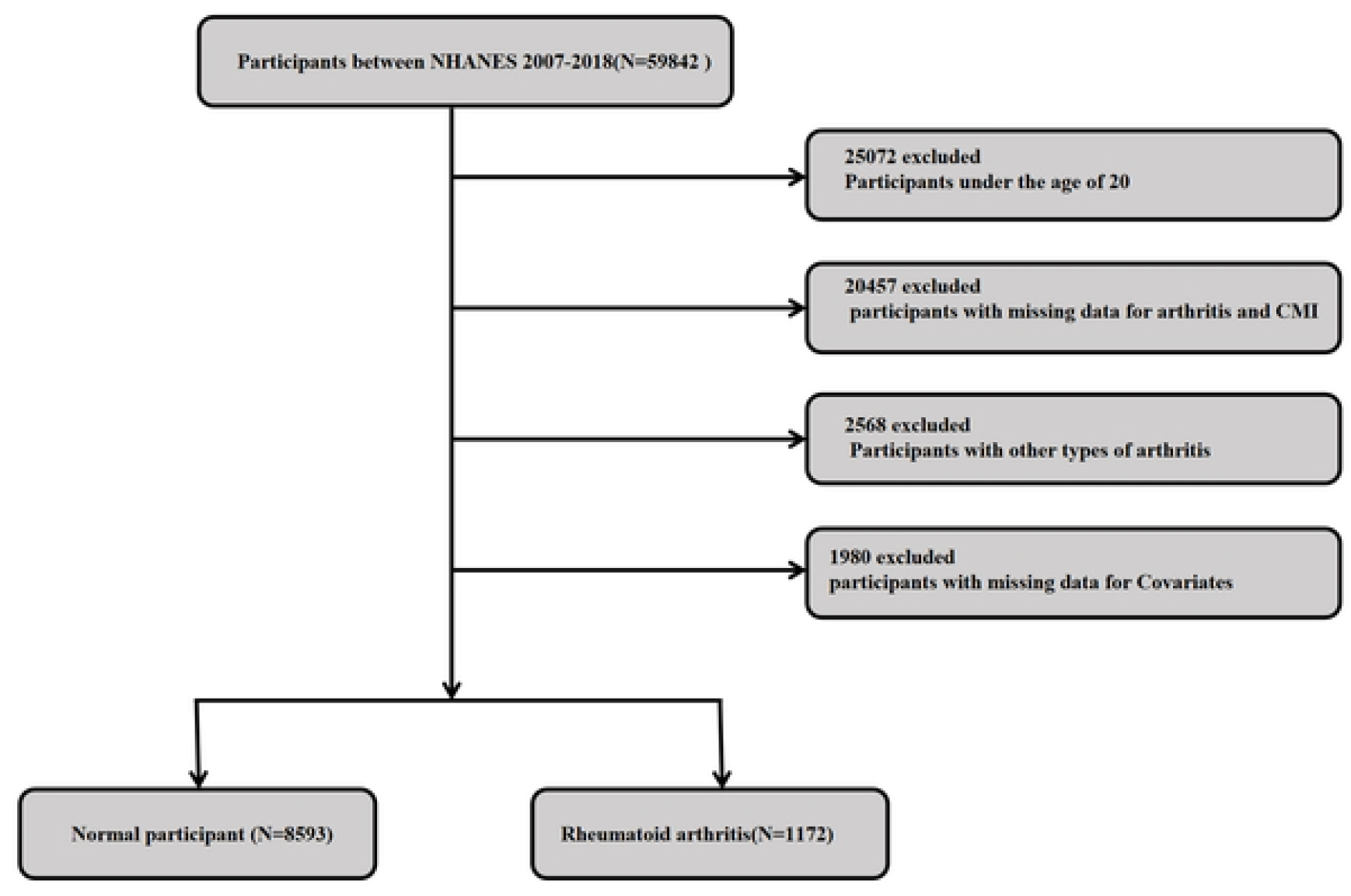
Flowchart of participant selection from NHANES 2007–2018.

### Exposure Variable: CMI Assessment

The cardiometabolic index (CMI) was calculated as follows [25]:

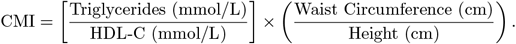

In this study, CMI was analyzed as a continuous exposure variable. Participants were also stratified into tertiles based on CMI values for subsequent analyses. The tertiles were defined as: T1: CMI *<* 0.34; T2: CMI 0.34–0.72; T3: CMI *>* 0.72.

### Outcome Variable: RA Assessment

RA was ascertained based on self-reported data obtained through personal interviews conducted as part of the health status questionnaire. Participants were first asked: “Has a doctor or other health professional ever told you that you have arthritis?” Those who responded “yes” were further asked: “What type of arthritis was it?” Individuals who reported “rheumatoid arthritis” were classified as having RA.

### Mediating Variables

Diabetes was defined as meeting any of the following criteria: (1) fasting plasma glucose ≥ 126 mg/dL; (2) glycated hemoglobin (HbA1c) *≥* 6.5%; (3) 2-hour oral glucose tolerance test (OGTT) result *≥*200 mg/dL (11.1 mmol/L); (4) current use of antidiabetic medication; or (5) a previous physician diagnosis. Hypertension status was determined based on averaged systolic (*≥*140 mmHg) and/or diastolic (*≥*90 mmHg) blood pressure measurements from the examination data, supplemented with self-reported physician diagnosis or current use of antihypertensive medication from the interview questionnaire. Furthermore, an extensive panel of laboratory parameters was collected, including neutrophil count, lymphocyte count, and platelet count. The following derived indices were computed: Neutrophil-to-Lymphocyte Ratio (NLR) = Neutrophil count / Lymphocyte count; Systemic Immune-Inflammation Index (SII) = Platelet count *×* Neutrophil count / Lymphocyte count.

### Covariates

This study incorporated covariates spanning demographic characteristics and lifestyle factors. Demographic variables included age, sex (male/female), race/ethnicity (Mexican American, non-Hispanic Black, non-Hispanic White, other Hispanic, other races), educational attainment (less than high school, high school or equivalent, college or above), and marital status (married or cohabiting with a partner, living alone). Lifestyle factors comprised the poverty-income ratio (PIR), smoking status (current smoker, former smoker, never smoker), alcohol consumption (yes/no), physical activity (active/inactive), and history of glucocorticoid medication use (yes/no). PIR was classified into three tiers: high (PIR ≥ 3.5), medium (1.3 ≤ PIR < 3.5), and low (PIR < 1.3) [26]. Current smokers were defined as individuals who had smoked more than 100 cigarettes in their lifetime and continued to smoke at the time of the survey. Former smokers were those who had smoked more than 100 cigarettes but were not current smokers. Never smokers were defined as participants who had smoked fewer than 100 cigarettes in their lifetime. Alcohol consumption was assessed using the following question: “During the past 12 months, have you consumed at least 12 drinks of any kind of alcoholic beverage?” A “drink” was defined as 12 ounces of beer, 4 ounces of wine, or 1 ounce of distilled spirits. Physical activity was evaluated via a self-reported questionnaire and quantified as metabolic equivalent task minutes per week (MET-min/week). Participants reporting no physical activity were assigned a value of zero. PA levels were dichotomized into “active” or “inactive” based on whether they met the U.S. physical activity guidelines, which recommend at least 150 minutes of moderate-intensity or 75 minutes of vigorous-intensity activity per week, corresponding to 600 MET-min/week [27]. Glucocorticoid use was identified through the question: “During the past 30 days, have you taken or used any prescription medication?” in conjunction with the review of medication names.

### Statistical Analysis

This cross-sectional study used data from the NHANES database to investigate the association between CMI and RA. In accordance with NHANES analytical guidelines for complex survey designs, appropriate sample weights, stratification variables, and clustering variables were applied to all analyses to ensure national representativeness. Study participants were categorized into low, medium, and high CMI groups according to tertiles. The normality of continuous variables was evaluated. Non-normally distributed continuous variables are summarized as weighted medians with interquartile ranges (IQRs), and group differences were examined using the Kruskal-Wallis H test, accounting for the survey design. Categorical variables are presented as unweighted counts (N) and weighted percentages (%), with comparisons performed via the Rao-Scott corrected chi-square test. Survey-weighted multivariable logistic regression models were used to assess the relationship between CMI and RA, modeling CMI both as a continuous variable and as a categorical variable (tertiles). We constructed three models with sequential adjustments: Model 1 was unadjusted; Model 2 adjusted for demographic characteristics (sex, age, race/ethnicity); Model 3 further adjusted for lifestyle and socioeconomic factors (marital status, education level, poverty-income ratio (PIR), alcohol use, smoking history, glucocorticoid use, and physical activity). Stratified analyses were conducted to examine potential effect modification by age, sex, race/ethnicity, education level, marital status, PIR, drinking status, smoking status, and physical activity level. Interaction terms were included in the models, and the significance of interactions was tested using likelihood ratio tests. To evaluate the nonlinear association between CMI and RA, we fitted restricted cubic splines (RCS) with four knots placed at the 5th, 35th, 65th, and 95th percentiles. If the overall test for nonlinearity yielded a P-value *<* 0.05, a two-piecewise linear regression model was applied to identify the threshold effect. The likelihood ratio test was used to compare the piecewise model with the linear model. To explore the potential pathways and statistically decompose the observed cross-sectional association between CMI and RA, we conducted an exploratory mediation analysis using a bootstrap approach with 2000 resamples to assess the following candidate mediators: hypertension, diabetes, and inflammatory markers—including neutrophil count, neutrophil-to-lymphocyte ratio (NLR), and the systemic immune-inflammation index (SII).Crucially, given the cross-sectional nature of the data, this analysis serves as a statistical description of how the association is partitioned and should not be interpreted as evidence of causal mediation. The proportion of the total association explained by each mediator was calculated.All statistical analyses were performed using R software (version 4.4.1). A P-value *<* 0.05 was considered statistically significant.

## Results

### Characteristics of Participants

Table1 summarizes the baseline characteristics of participants categorized into tertiles based on the Cardiometabolic Index (CMI): T1 (low CMI, *n* = 3255), T2 (medium CMI, *n* = 3255), and T3 (high CMI, *n* = 3255). A graded increase was observed in the prevalence of rheumatoid arthritis (RA) (T1: 9.74%; T2: 12.28%; T3: 15.81%; *P <* 0.001), hypertension (T1: 22.43%; T2: 34.61%; T3: 46.85%; *P <* 0.001), and diabetes (T1: 5.76%; T2: 13.12%; T3: 27.02%; *P <* 0.001) across ascending CMI tertiles. Additionally, higher CMI levels were associated with a greater proportion of males, older age, and a higher representation of non-Hispanic white individuals. These participants also exhibited lower educational attainment, reduced household income, higher current smoking rates, lower physical activity levels, and elevated systemic inflammatory markers, including neutrophil count, SII, and NLR. All reported differences were statistically significant.

**Table 1.** Weighted features of the study population.

| Characteristic | Total (N = 9765) | CMI tertile |  |  | P value |
| --- | --- | --- | --- | --- | --- |
|  |  | T1(n=3255) | T2(n=3255) | T3(n=3255) |  |
| Weighted No. | 71294057 | 24704928 | 23522316 | 23066813 |  |
| Age (median, IQR) | 45.00 (31.00, 58.00) | 41.00 (28.00, 55.00) | 45.00 (32.00, 59.00) | 47.00 (35.41, 60.00) | < 0.001 |
| **Sex, n (%)** |  |  |  |  |  |
| Male | 4950(49.47) | 1320(40.03) | 1641(50.46) | 1989(61.86) | < 0.001 |
| Female | 4185(50.53) | 1935(59.97) | 1614(49.54) | 1266(38.14) |  |
| **Race/ethnicity, n(%)** |  |  |  |  |  |
| Mexican American | 1545(8.99) | 329(6.01) | 538(9.19) | 678(11.97) | < 0.001 |
| Non - Hispanic Black | 1864(9.79) | 870(13.7) | 646(9.98) | 348(5.40) |  |
| Non - Hispanic White | 4131(67.88) | 1322(67.41) | 1336(67.38) | 1473(68.9) |  |
| Other Hispanic | 1017(5.72) | 260(4.73) | 344(5.98) | 413(6.51) |  |
| Other Race | 1208(7.62) | 474(8.15) | 391(7.46) | 343(7.21) |  |
| **Education level, n(%)** |  |  |  |  |  |
| <High school diploma | 2159(14.21) | 509(10.00) | 767(15.15) | 883(17.75) | < 0.001 |
| High school diploma | 2164(22.29) | 658(19.89) | 720(21.14) | 786(25.85) |  |
| College diploma | 5442(63.50) | 2088(70.11) | 1768(63.52) | 1586(56.41) |  |
| **Marital status, n(%)** |  |  |  |  |  |
| Married/living with partner | 5876(63.67) | 1803(61.41) | 2001(64.34) | 2072(65.43) | 0.035 |
| Living alone | 3889(36.33) | 1452(38.59) | 1254(35.66) | 1183(34.57) |  |
| **PIR, n(%)** |  |  |  |  |  |
| Low | 3023(21.15) | 891(18.67) | 991(20.63) | 1141(24.31) | < 0.001 |
| Medium | 3732(35.88) | 1232(33.88) | 1250(36.17) | 1250(37.71) |  |
| High | 1950(18.73) | 1132(47.44) | 1014(43.19) | 864(37.97) |  |
| **Alcohol drinks, n (%)** |  |  |  |  |  |
| Yes | 7468(81.35) | 2529(82.48) | 2442(80.49) | 2497(81.01) | 0.204 |
| No | 2297(18.65) | 726(17.52) | 813(19.51) | 758(18.99) |  |
| **Smoke, n(%)** |  |  |  |  |  |
| Never | 5576(57.28) | 2078(63.23) | 1869(57.65) | 1629(50.54) | < 0.001 |
| Former | 2239(23.99) | 605(20.82) | 750(23.53) | 884(27.87) |  |
| Current | 1950(18.73) | 572(15.96) | 636(18.83) | 742(21.59) |  |
| **Physical activity, n(%)** |  |  |  |  |  |
| active | 6604(71.70) | 2408(77.42) | 2160(71.13) | 2036(66.14) | < 0.001 |
| inactive | 3161(28.30) | 847(22.58) | 1095(28.87) | 1219(33.86) |  |
| **history of glucocorticoid use, n(%)** |  |  |  |  |  |
| Yes | 104(0.94) | 31(0.87) | 31(0.76) | 42(1.18) | 0.39 |
| No | 9661(99.06) | 3224(99.13) | 3224(99.24) | 3213(98.82) |  |
| **Hypertension (%)** |  |  |  |  |  |
| Yes | 3695(34.35) | 867(22.43) | 1262(34.61) | 1566(46.85) | < 0.001 |
| No | 6070(65.65) | 2388(77.57) | 1993(65.39) | 1689(53.15) |  |
| **Diabetes, n(%)** |  |  |  |  |  |
| Yes | 1924(15.07) | 286(5.76) | 587(13.13) | 1051(27.03) | < 0.001 |
| No | 7841(84.93) | 2969(94.24) | 2668(86.87) | 2204(72.97) |  |
| **RA, n(%)** |  |  |  |  |  |
| Yes | 1172(12.54) | 302(9.74) | 385(12.28) | 485(15.81) | < 0.001 |
| No | 8593(87.46) | 2953(90.26) | 2870(87.72) | 2770(84.19) |  |
| **Neutrophil (median, IQR)** |  |  |  |  |  |
|  | 3.70 (2.90, 4.70) | 3.20 (2.50, 4.10) | 3.80 (2.90, 4.80) | 4.10 (3.30, 5.11) | < 0.001 |
| **SII (median, IQR)** |  |  |  |  |  |
|  | 443.67 (322.74, 623.50) | 401.34 (295.95, 570.40) | 463.34 (336.44, 636.93) | 470.59 (344.72, 651.55) | < 0.001 |
| **NLR (median, IQR)** |  |  |  |  |  |
|  | 1.91 (1.46, 2.53) | 1.79 (1.40, 2.38) | 1.95 (1.48, 2.56) | 2.00 (1.52, 2.62) | < 0.001 |

Table notes: RA, rheumatoid arthritis; CMI, cardiometabolic index; PIR, Family Income to Poverty Ratio; SII, Systemic Immune-Inflammation Index; NLR, Neutrophil-to-Lymphocyte Ratio.

### Association Between CMI and RA

As summarized in Table 2, a significant positive association was observed between CMI and RA across all regression models. Specifically, elevated CMI was associated with increased odds of RA in the unadjusted model (OR = 1.14, 95% CI: 1.05–1.24), the partially adjusted model (OR = 1.20, 95% CI: 1.09–1.33), and the fully adjusted model (OR = 1.20, 95% CI: 1.08–1.33). This association persisted when CMI was analyzed in tertiles, with a significant trend toward higher RA prevalence across increasing tertiles (*P* for trend *<* 0.001). In the fully adjusted model, individuals in the highest CMI tertile exhibited a 74% greater risk of RA compared to those in the lowest tertile (OR = 1.74, 95% CI: 1.33–2.28). Moreover, restricted cubic spline (RCS) regression revealed a nonlinear relationship between CMI and RA (Figure2), with an inflection point at 1.63. A two-piecewise linear regression model further illustrated a pronounced association below the threshold (CMI *<* 1.63; OR = 1.53, 95% CI: 1.27–1.85), whereas above this value (CMI *≥*1.63), the association was attenuated and not statistically significant (OR= 1.07, 95% CI: 0.97–1.18), as detailed in Table3.

**Table 2.** Weighted multivariable logistic regression analysis of the association between CMI and RA.

| Variables | Model 1 |  | Model 2 |  | Model 3 |  |
| --- | --- | --- | --- | --- | --- | --- |
|  | OR (95%CI) | p-value | OR (95%CI) | p-value | OR (95%CI) | p-value |
| CMI |  |  |  |  |  |  |
| Continuous | 1.14 (1.05, 1.24) | 0.002 | 1.20 (1.09, 1.33) | < 0.001 | 1.20 (1.08, 1.33) | < 0.001 |
| CMI (quartiles) |  |  |  |  |  |  |
| T1 | Ref |  | Ref |  | Ref |  |
| T2 | 1.30 (1.03, 1.64) | 0.030 | 1.22 (0.94, 1.57) | 0.131 | 1.24 (0.95, 1.61) | 0.106 |
| T3 | 1.74 (1.37, 2.20) | < 0.001 | 1.72 (1.34, 2.22) | < 0.001 | 1.74 (1.33, 2.28) | < 0.001 |
| P for trend | < 0.001 |  | < 0.001 |  | < 0.001 |  |

Table notes: OR, Odds Ratio; CI, Confidence Interval. Model 1: Crude; Model 2: adjusted for sex, age, race/ethnicity; Model 3: further adjusted for marital status, education level, PIR, alcohol use, smoking history, glucocorticoid use, and physical activity.

**Table 3.** Threshold effect analysis of the link between CMI and RA.

| Variables | OR (95%CI) | p-value |
| --- | --- | --- |
| Inflection Point | 1.63 (1.27,1.85) | < 0.001 |
| < Inflection Point | 1.53 (1.27,1.85) | < 0.175 |
| > Inflection Point | 1.07 (0.97,1.18) | 0.175 |
| Likelihood Ratio Test |  | 0.001 |

Table notes: Adjusted for confounders specified in Model 3.

**Fig 2.**
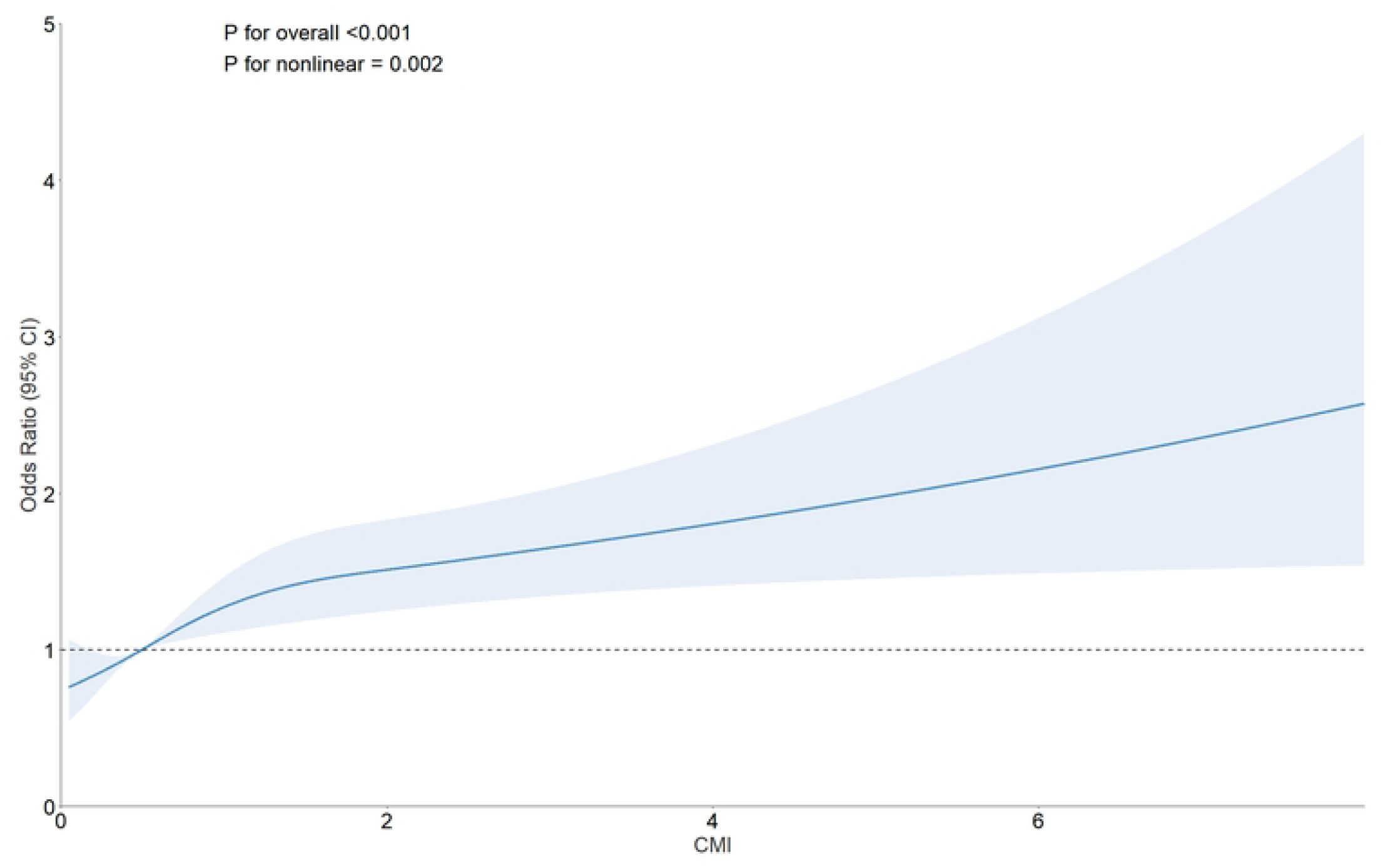
Restricted cubic spline curve (RCS) plot of the relationship between CMI and RA incidence. Adjusted for confounders specified in Model 3.

### Subgroup Analysis

To assess the robustness of the association between CMI and rheumatoid arthritis (RA), we performed subgroup analyses and interaction tests across multiple covariates, including age, sex, race, education level, marital status, PIR, alcohol consumption, smoking status, and physical activity level. As illustrated in Figure3, a significant positive association was observed between CMI and RA, which remained consistent across nearly all subgroups. Of note, the interaction test for age stratification indicated a marginally significant moderating effect (*P* for interaction = 0.054), suggesting that age may play a potential role in modulating the relationship between CMI and RA. However, this finding did not meet the conventional threshold for statistical significance (*P <* 0.05), and its clinical relevance warrants further investigation. For all other subgroups, interaction P-values exceeded 0.05, demonstrating a consistent association between CMI and RA across diverse demographic and lifestyle strata, with no evidence of significant heterogeneity.

**Fig 3.**
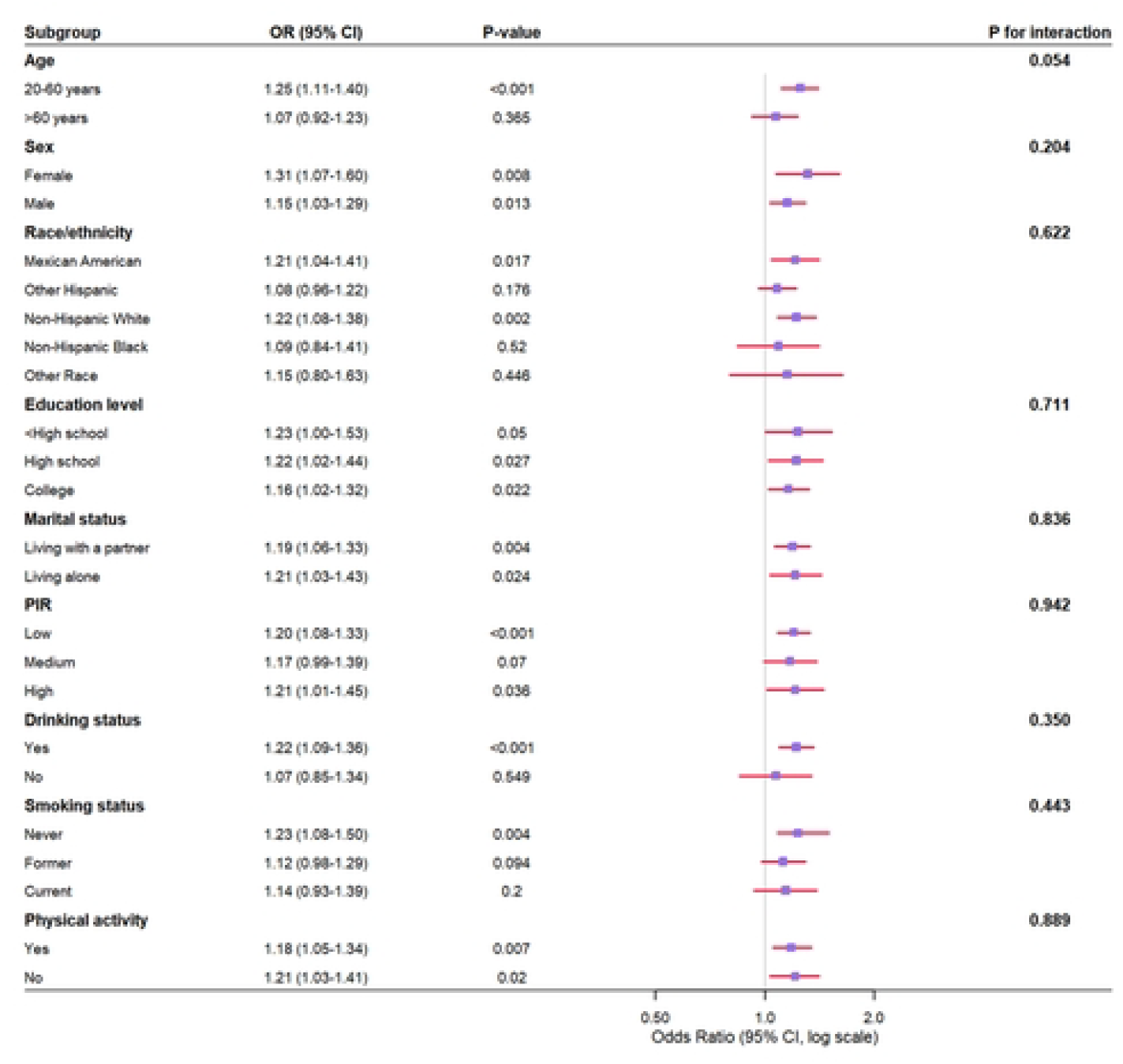
Association of CMI with RA across different subgroups. Adjusted for confounders specified in Model 3. However, any variable utilized as a stratification factor was excluded from the adjustment.

### Exploratory Mediation Analysis

An exploratory mediation analysis was performed to investigatethe extent to which the observed association between CMI and RA could be statistically explained by hypertension, diabetes, and inflammatory markers. As depicted in Figure4, hypertension, diabetes, and neutrophil count were significant explanatory variables in this relationship, with estimated indirect effect values of 0.0033 (*P <* 0.001), 0.0024 (*P* = 0.001), and 0.0008 (*P <* 0.001), respectively. The proportions of the total association statistically explained by these variables were 22.2%, 15.9%, and 5.4%, respectively. In contrast, neither SII nor NLR demonstrated significant explanatory roles.

**Fig 4.**
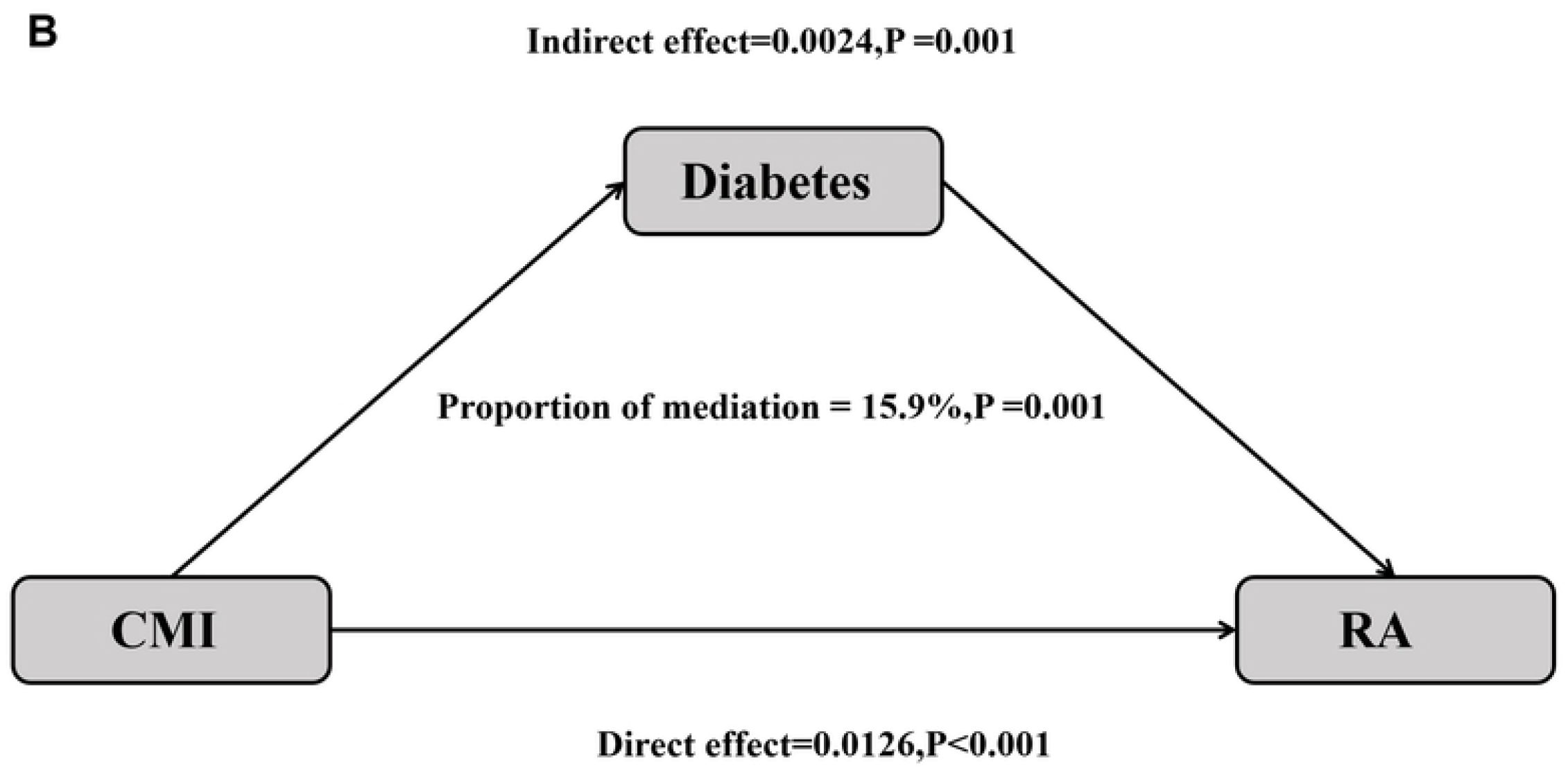
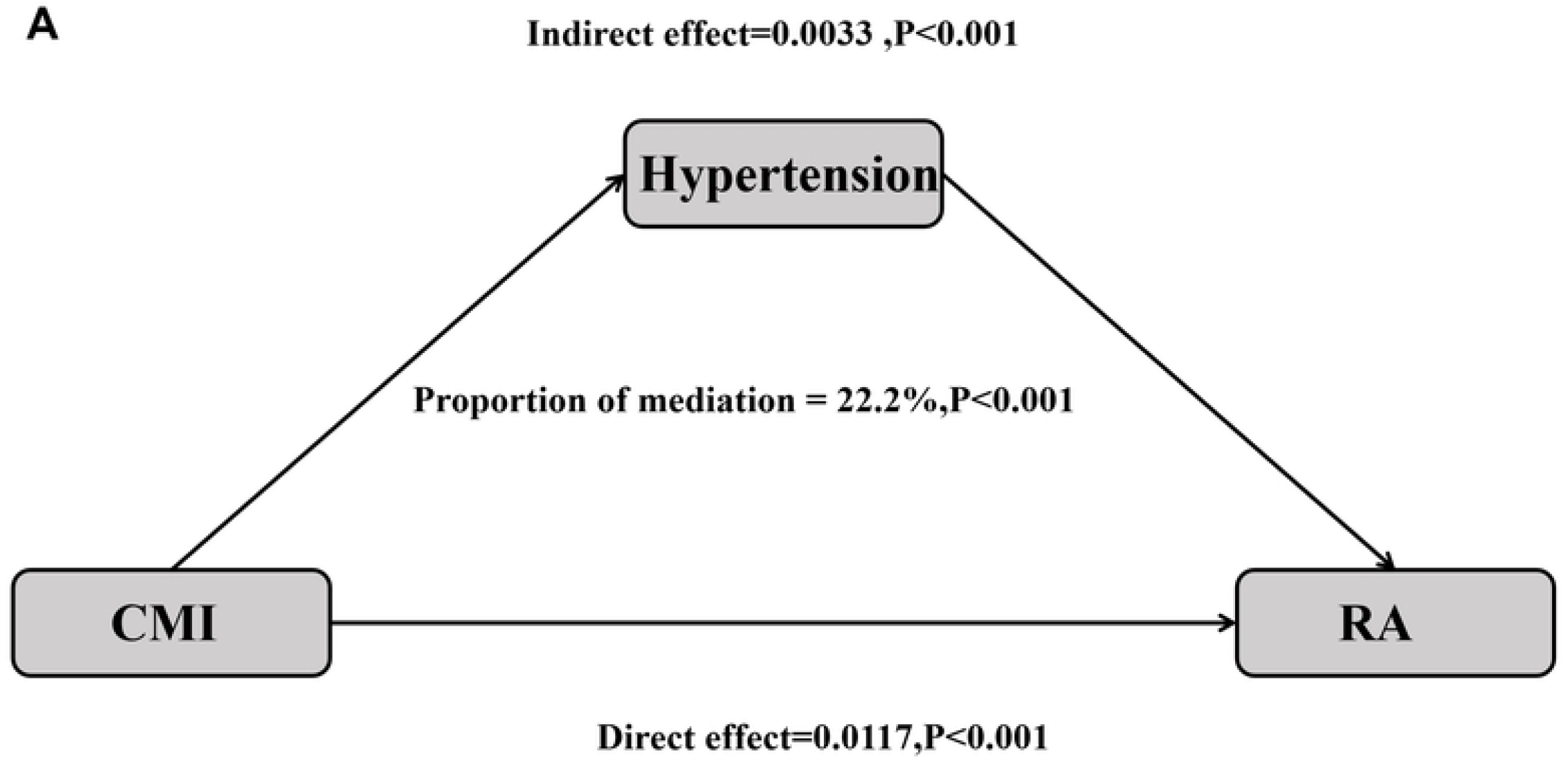
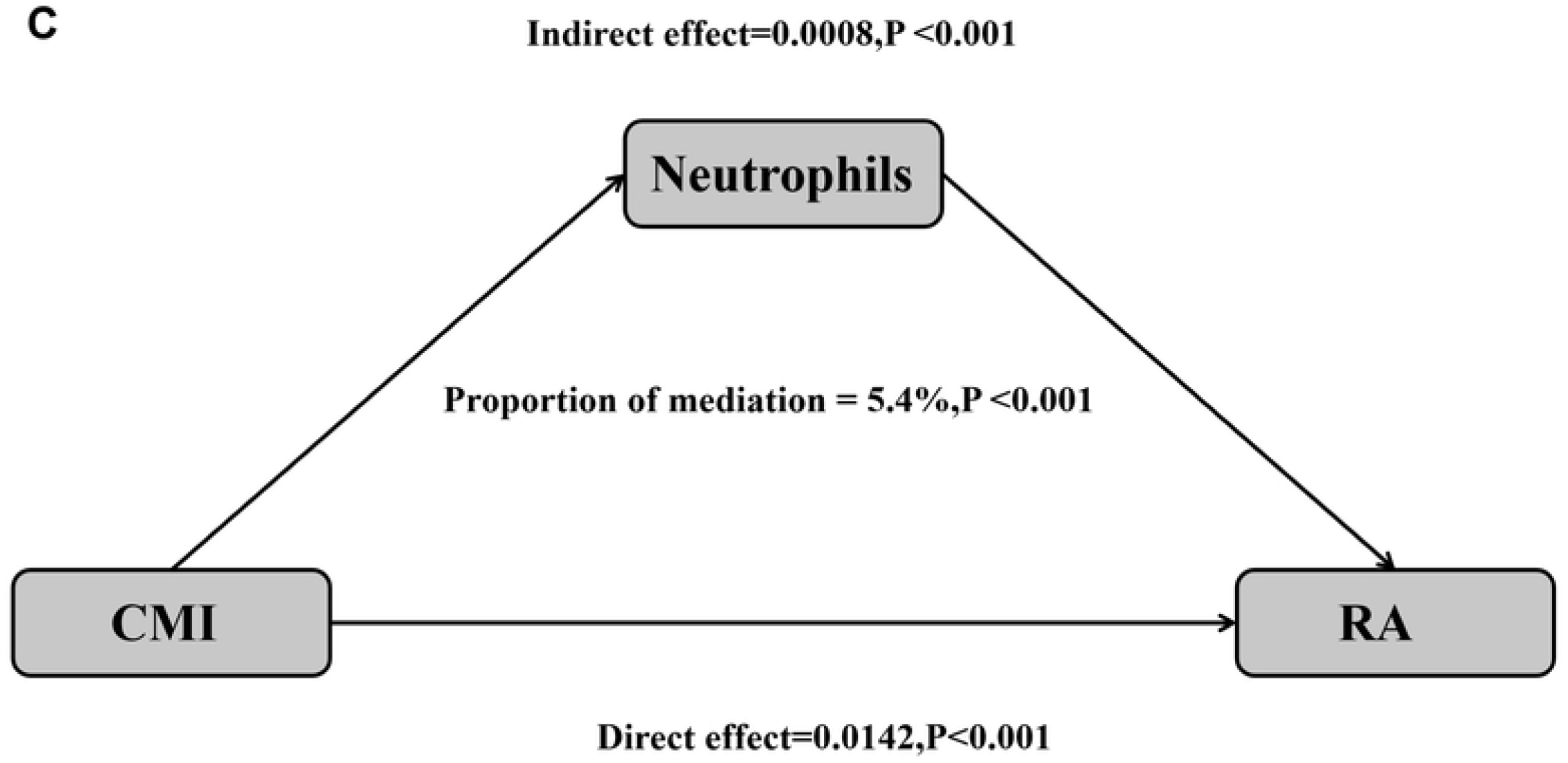
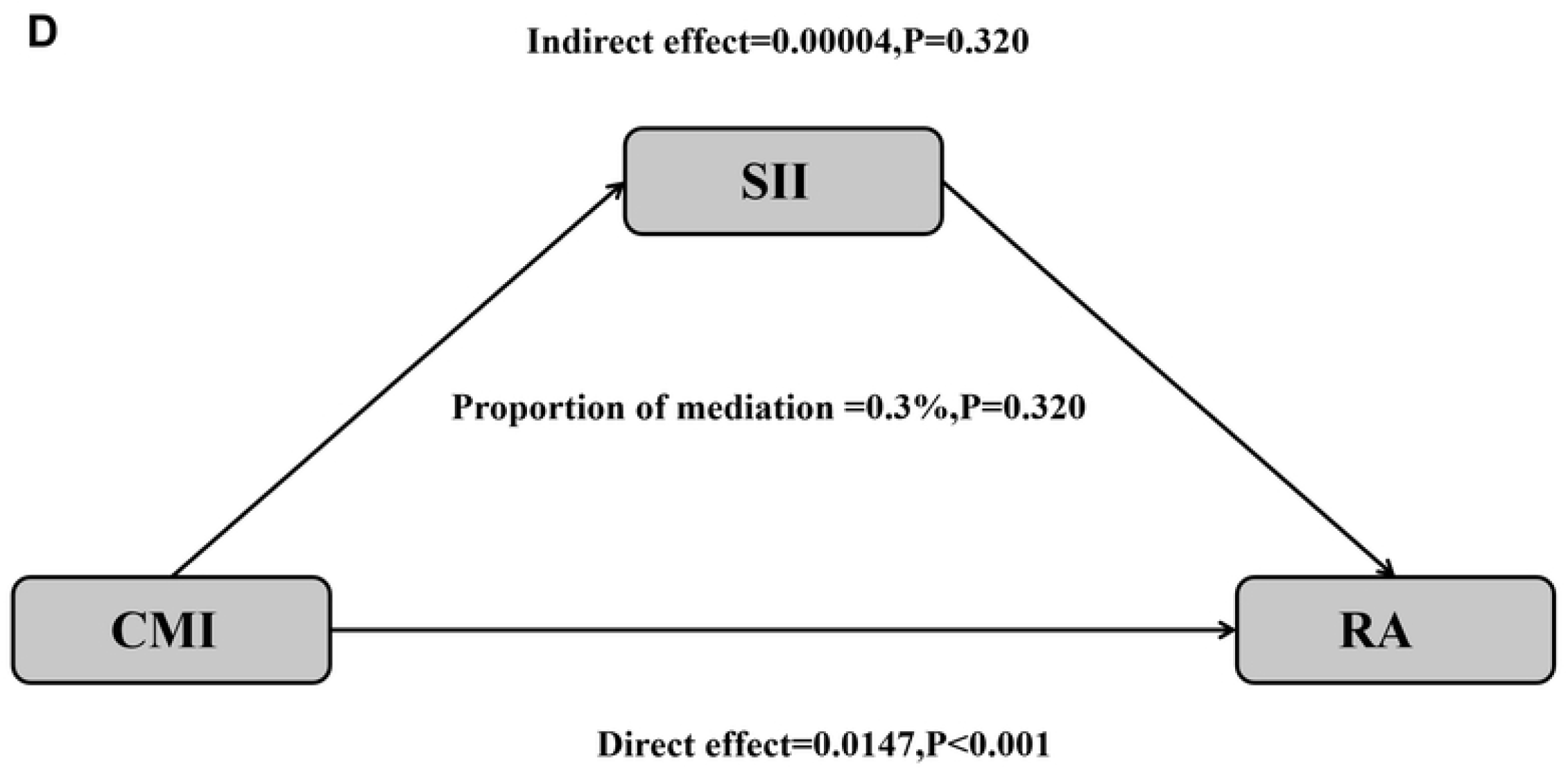
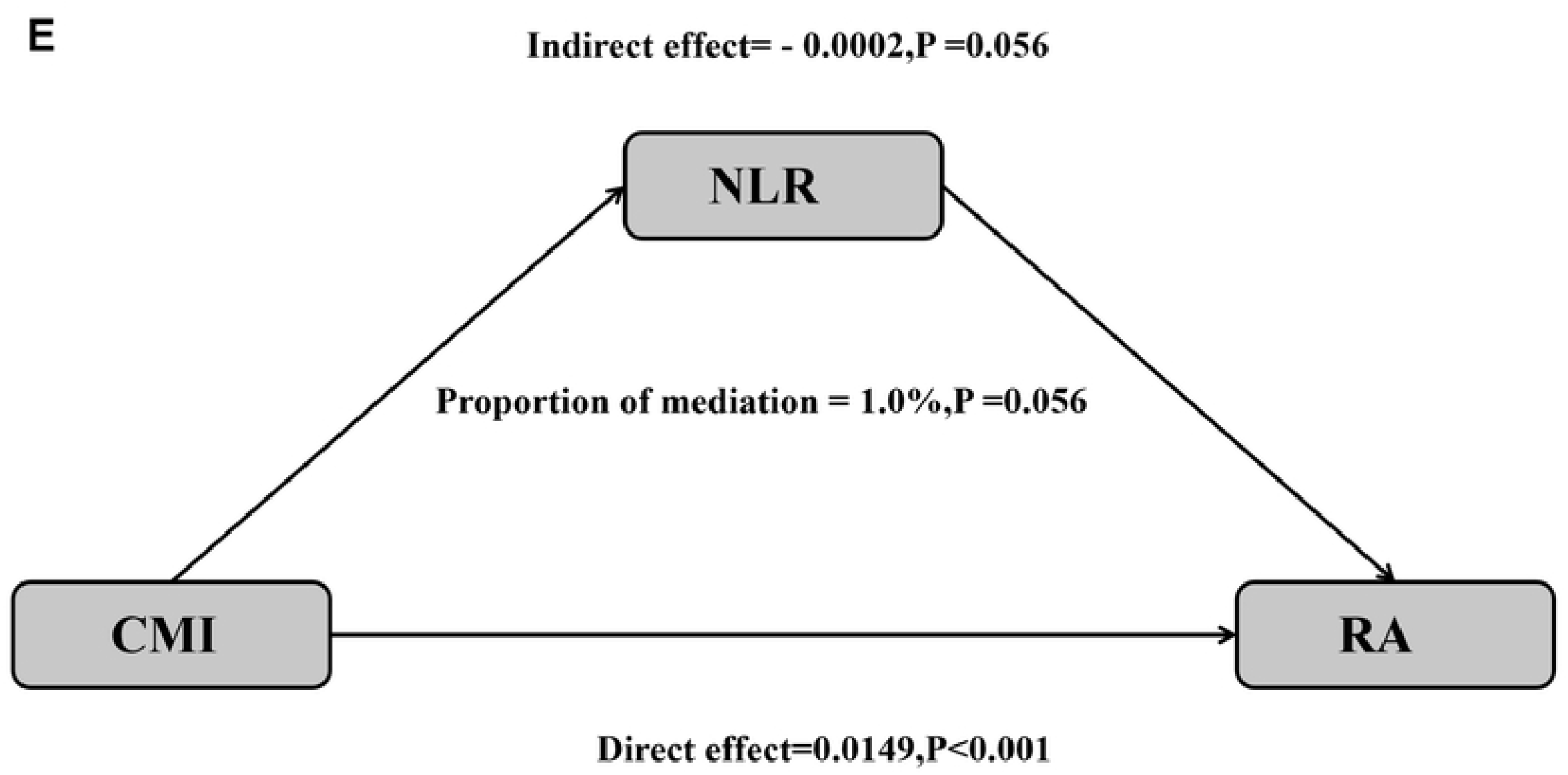
Exploratory Mediation Analysis. Analysis of the mediating effects of hypertension (A), diabetes (B), neutrophil count (C), SII (D), and NLR (E) on the relationship between CMI and RA. Adjusted for confounders specified in Model 3.

## Discussion

This study provides the first systematic assessment of the association between the cardiometabolic index (CMI) and the prevalence of RA. In a large-scale cross-sectional analysis of 9,765 U.S. adults (weighted *N* = 71, 294, 057), we demonstrated that higher CMI levels are independently and significantly associated with an increased prevalence of RA, exhibiting a clear dose-response relationship, even after comprehensive adjustment for sociodemographic, lifestyle, and clinical confounders. Specifically, individuals in the highest CMI tertile exhibited a 74% increased risk of RA (OR = 1.74, 95% CI: 1.33–2.28) compared to those in the lowest tertile. Notably, restricted cubic spline analysis revealed a nonlinear relationship characterized by a clear threshold effect. Below the CMI value of 1.63, the association with RA risk was pronounced (OR = 1.53, 95% CI: 1.27–1.85). Beyond this inflection point, however, the association lost statistical significance. This nonlinear pattern represents a key finding, suggesting that CMI may serve as a sensitive marker within a specific range in the pathophysiology of RA. Consistent associations were observed across all predefined subgroups without significant heterogeneity. Our analysis of potential pathways revealed that a substantial portion of the association between CMI and RA was mediated by hypertension and diabetes, whereas neutrophil count contributed only minimally to the mediating effect.The direct effect of CMI on RA remained highly significant (*P <* 0.001) in all mediation models. In conclusion, our findings not only strengthen the evidence linking cardiometabolic dysfunction to RA but also introduce CMI as a novel and practical tool for improving risk stratification and identifying individuals at high risk for RA.

Our conclusions align with the evolving understanding of metabolic dysregulation as a contributor to RA risk. The French E3N Cohort Study reported a significant association between abdominal obesity (waist circumference *>* 88 cm) and incident RA (HR = 1.20, 95% CI: 1.00–1.50), independent of BMI [28]. Similarly, a Swedish nested case-control study by Lotta et al. found positive correlations for both BMI and waist circumference with RA risk—a 5 kg/m^2^ increase in BMI corresponded to a 13% higher risk (OR = 1.13, 95% CI: 1.00–1.28), and each 1 cm increase in waist circumference was associated with a 2% rise in risk (OR = 1.02, 95% CI: 1.01–1.04)—firming support for abdominal obesity as an independent risk factor [29]. This trend is echoed in female-specific research; a prospective cohort study from Harvard Medical School found that women maintaining an overweight (25 *≤* BMI *<* 30 kg/m^2^) or obese (BMI *≥* 30 kg/m^2^) status faced a significantly elevated future risk of RA (Obese: HR = 1.23, 95% CI: 1.06–1.44; Overweight: HR = 1.34, 95% CI: 1.06–1.68) [30]. Expanding on this, Luo et al., in a large prospective cohort of 369,065 participants, demonstrated that metabolic syndrome (MetS) and four of its individual components were associated with increased RA risk: the adjusted HR for MetS was 1.22 (95% CI: 1.14–1.30). Significant associations were also observed for elevated waist circumference (HR = 1.21, 95% CI: 1.12–1.32), high triglycerides (HR = 1.12, 95% CI: 1.05–1.19), low HDL-C (HR = 1.31, 95% CI: 1.23–1.39), and hyperglycemia (HR = 1.15, 95% CI: 1.05–1.25). Moreover, RA risk escalated with the number of MetS components present, reaching its peak among individuals exhibiting all five components [31]. However, prior research has largely relied on isolated metrics of lipid metabolism or adiposity, limiting a holistic view of the metabolic-RA relationship. Our study is the first to employ CMI—a composite metric integrating central obesity (waist-to-height ratio) and atherogenic dyslipidemia (TG/HDL-C ratio)—in RA association analysis. We not only establish a significant positive correlation between CMI and RA risk in a U.S. population but also uncover a nuanced nonlinear relationship characterized by a threshold effect. This insight offers a novel theoretical foundation and suggests that CMI could be a potential marker for identifying individuals at risk, a hypothesis that needs to be tested in future interventional studies.

The cardiometabolic index (CMI) incorporates two core components—waist-to-height ratio (WHtR) and the triglyceride to high-density lipoprotein cholesterol (TG/HDL-C) ratio—which together elucidate its association with RA through distinct yet complementary pathological pathways. WHtR is a sensitive surrogate for visceral adipose tissue (VAT) accumulation [32]. VAT is not an inert energy store but a dynamic endocrine and immunologic organ that secretes a plethora of bioactive molecules termed adipokines [33]. In obesity, VAT undergoes functional dysregulation, shifting its secretory profile towards a pro-inflammatory phenotype. This involves increased secretion of pro-inflammatory adipokines like TNF-*α*, IL-6, and leptin—key drivers of synovial inflammation and joint destruction in RA that directly promote immune cell activation, proliferation, differentiation, and osteoclastogenesis [34, 35]. Consequently, chronically elevated levels of these cytokines in high-CMI individuals create a “primed” systemic pro-inflammatory microenvironment, facilitating the initiation of autoimmune responses. Concurrently, secretion of anti-inflammatory adipokines, notably adiponectin which suppresses inflammation and enhances insulin sensitivity, is diminished [36, 37]. This reduction further compromises the body’s ability to regulate inflammation, exacerbating the imbalance between pro- and anti-inflammatory forces. This VAT-driven chronic low-grade systemic inflammation can persistently erode immune tolerance, thereby providing a pathological backdrop for autoimmunity against joint antigens.The TG/HDL-C ratio, the second component of CMI, is a well-established marker of insulin resistance (IR) [38]. IR itself exerts potent pro-inflammatory effects beyond its role in metabolic dysregulation. Hyperinsulinemia and hyperglycemia can activate innate immune cells (e.g., monocytes/macrophages) via signaling pathways such as NF-*κ*B and mTOR, potentiating their pro-inflammatory cytokine output [39, 40]. Furthermore, the IR milieu catalyzes “immunometabolic reprogramming” in adaptive immune cells like T and B lymphocytes. For instance, effector T cells (Th1, Th17), pivotal in RA pathogenesis, are highly dependent on glycolytic metabolism for their activation and expansion. The metabolic environment characteristic of IR supports and promotes this glycolytic switch, thereby driving the pathogenic activation and proliferation of Th1/Th17 cells and worsening joint inflammation [41, 42]. Additionally, the atherogenic dyslipidemia (high TG, low HDL-C) captured by this ratio contributes directly to immune dysregulation. Oxidatively modified lipoproteins can act as damage-associated molecular patterns (DAMPs), triggering inflammatory responses upon recognition by immune cells [43, 44]. Conversely, reduced levels of HDL, which possesses anti-inflammatory, antioxidant, and cholesterol efflux capabilities, impair the clearance of inflammatory stimuli and hinder tissue repair. In essence, the IR and dyslipidemia reflected by an elevated TG/HDL-C ratio provide both aberrant metabolic signals (“erroneous fuel”) and dysregulated immune instructions, collectively propelling the immune system from homeostasis towards a pathological state of autoimmunity targeting the joints.

Nonlinear regression identified a significant threshold effect in the relationship between CMI and RA risk, with an inflection point at 1.63. Below this threshold (CMI *<* 1.63), each unit increase in CMI was associated with a 53% higher risk of RA (OR = 1.53, 95% CI: 1.27–1.85). Beyond this level, the association was no longer statistically significant. This pattern closely aligns with the biological properties of CMI: at lower values, increases in CMI sensitively capture worsening early metabolic dysregulation and exhibit a clear dose-response relationship with RA risk. At higher CMI levels, the attenuated association may reflect a “ceiling effect” where adipose tissue dysfunction reaches a plateau in its capacity to drive further pro-inflammatory changes relevant to RA pathogenesis. Alternatively, individuals with very high CMI are more likely to be on pharmacological interventions (e.g., statins, antihypertensives), which themselves can modulate inflammation and RA risk, potentially confounding the association. [45–47].Exploratory mediation analysis suggested that hypertension (explaining 22.2% of the total association) and diabetes (15.9%) were the primary factors statistically underlying the CMI–RA association, whereas neutrophils showed a relatively modest mediating effect (5.4%). These results suggest that metabolic dysregulation, as captured by CMI, contributes to RA risk largely through the development of core cardiometabolic comorbidities—rather than via direct biochemical pathways. Such comorbidities promote a pro-autoimmune “pathogenic soil” through systemic alterations including vascular endothelial dysfunction, oxidative stress, and accumulation of advanced glycation end products (AGEs) [48]. Moreover, these metabolic conditions and RA share numerous inflammatory pathways and risk factors, potentially initiating a self-reinforcing cycle that amplifies the detrimental impact of metabolic dysregulation on immune homeostasis [49, 50]. Given that these findings are derived from a cross-sectional study design, they should be interpreted as generating hypotheses about potential mechanisms rather than confirming causal mediation. Further validation in prospective cohort studies is still required.

## Limitations

This study has several limitations. First, due to its cross-sectional design, no causal inference can be established between CMI and RA. Future longitudinal cohort studies or interventional trials are warranted to clarify the potential role of CMI in the pathogenesis and progression of RA.Second, although we adjusted for a range of known confounders based on existing literature and clinical expertise, residual confounding may still be present. Unmeasured factors, such as disease activity and severity of RA, dietary habits, and specific nutrient intakes, could influence the observed associations.Third, the definition of RA was based on self-reported physician diagnosis rather than clinical examination or classification criteria (e.g., ACR/EULAR criteria). While this method is specific for confirming diagnosed cases, it lacks sensitivity and may miss individuals with undiagnosed or early-stage disease. This non-differential misclassification could potentially bias our effect estimates towards the null, meaning the true strength of the association between CMI and RA might be even stronger than what we observed. Future studies utilizing clinically validated RA outcomes are warranted to confirm our findings.

## Conclusion

Our analysis revealed a significant positive association between CMI and the prevalence of RA among U.S. adults, characterized by a nonlinear threshold effect. Elevated CMI levels were significantly associated with an increased risk of RA. Specifically, for individuals with a CMI below 1.63, each unit increase in CMI corresponded to a 53% higher risk of RA.Notably, hypertension and diabetes were identified as key mediators in this relationship, accounting for 22.2% and 15.9% of the observed association, respectively. Although neutrophil count also demonstrated a significant mediating effect, its contribution was modest (proportion mediated: 5.4%).These findings offer novel mechanistic insights into the link between CMI and RA. Consequently, integrated management of cardiometabolic health may represent a critical preventive strategy for RA.

## Data Availability

All data used in this study are from publicly available sources and are freely accessible and available for use. For more information, please visit the following website:https://wwwn.cdc.gov/Nchs/ Nhanes/.

https://wwwn.cdc.gov/Nchs/Nhanes/

## Ethics approval and consent to participate

The data used in this analysis were accessed and downloaded from the NHANES website (https://wwwn.cdc.gov/nchs/nhanes/) on February 19, 2025. Given that the public-use datasets of NHANES have undergone rigorous de-identification and contain no direct personal identifiers (e.g., names, addresses, full dates of birth), the authors had no access to information that could identify individual participants at any stage of this study. Additionally, the NHANES study protocol has been approved by the Research Ethics Review Board of the National Center for Health Statistics (NCHS), and all participants have signed written informed consent forms.

## Consent for publication

Not applicable. This manuscript does not contain any individual person’s data in any form.

## Acknowledgments

The authors thank the National Center for Health Statistics (NCHS) for making the NHANES data publicly available, as well as all participants and staff of NHANES for their valuable contributions.

